# A tool for assessing changes in food preferences and health perceptions during nutritional interventions

**DOI:** 10.64898/2026.05.06.26352307

**Authors:** Maya Bar Or, Nuphar Vinegrad, Sara Menashe Auman, Idit F Liberty, Tom Schonberg

## Abstract

Understanding how nutritional interventions alter food evaluations may help clarify mechanisms of dietary behavior change; however, most studies focus on intake outcomes and rarely assess within-person changes in subjective food evaluation. We developed a brief, image-based rating tool that measures two core dimensions of food evaluation, liking and perceived healthiness, using standardized food images.

The tool was piloted in adults with type 2 diabetes participating in a medically supervised intervention that included structured glucose monitoring and professional dietary guidance. Ratings were collected at baseline, post-monitoring, and follow-up. In line with the methodological aim of this study, we examined whether the tool demonstrates internal coherence, sensitivity to change, and external validity against expert ratings and physiological measures, and whether it can capture item-level patterns relevant to eating behavior.

Results provide preliminary evidence that the tool is feasible, it is low-burden, and capable of detecting coherent relationships between food liking and health perceptions, including coordinated within-person changes over time and meaningful associations with external benchmarks. To support scalability and self-administration, we also developed an online smartphone-based demonstration version to exemplify the task structure and user experience. Overall, this pilot study suggests that a short, flexible rating task can serve as a practical measurement tool for tracking intervention-relevant changes in food evaluation and for informing the design of future nutritional interventions.

## Introduction

Healthy eating plays a crucial role in preventing chronic diseases such as obesity, cardiovascular disease, and diabetes. Yet achieving a balanced diet remains challenging in many societies (Bédard et al., 2020; Schwingshackl et al., 2017; Werle et al., 2013). Understanding how nutritional interventions change food preferences and health-related food beliefs is therefore critical for improving dietary behavior. Although many interventions evaluate dietary outcomes using intake-based metrics (e.g., food frequency questionnaires, dietary recalls, adherence indices), these approaches often do not directly assess changes in subjective food evaluation, mainly, how appealing foods are perceived to be (“liking”) and how appropriate they are judged for health goals (“healthiness”) (Burch et al., 2018; Schwingshackl et al., 2017). Capturing these perceptual changes is important, as shifts in beliefs and preferences may represent intermediate psychological mechanisms through which interventions exert long-term effects on eating behavior.

A balanced diet is a key component of a healthy lifestyle, and public health initiatives often promote nutritious food consumption. However, their effectiveness can be undermined by ingrained cultural perceptions and individual preferences (Schwingshackl et al., 2017). One prominent barrier is the widespread belief that healthy food is less palatable. This association has been extensively documented in studies of the “unhealthy = tasty” intuition, which is particularly prevalent in Western cultures (Haasova & Florack, 2019; Raghunathan et al., 2006; Werle et al., 2013). Recent research, including intervention-based studies, provided compelling evidence that altering health perceptions can directly influence food liking. For example, interventions that reframe healthy foods as pleasurable and emphasize their sensory attributes have been shown to increase both their perceived tastiness and actual preference among patients (Bédard et al., 2020; Haasova & Florack, 2019; Nadricka et al., 2020).

Evidence from cross-cultural studies further supports these findings. For instance, research conducted in France indicated that individuals are more likely to enjoy and consume nutritious foods in contexts where healthiness and tastiness are perceived as compatible (Werle et al., 2013). Moreover, targeted interventions that leverage this compatibility have been shown to effectively shift food preferences. In particular, patient sensory-focused approaches that emphasize pleasurable attributes of healthy foods, such as texture, flavor, and visual appeal, were associated with higher enjoyment and greater willingness to incorporate these foods into regular diet patterns (Bédard et al., 2020; Vogel & Mol, 2014). Together, these findings underscore the value of evidence-based strategies aimed at narrowing the perception gap between healthiness and tastiness.

In light of this evidence, there is a clear need for empirical tools that can sensitively track changes in food preferences and health-related beliefs in response to nutritional interventions. Although such interventions may help individuals form clearer links between dietary behavior and physiological outcomes, existing methodologies rarely assess how these experiences alter subjective food perception.

Here, we introduce and pilot a brief, computer-based tool designed to quantify changes in food evaluation during health-related interventions. The tool uses standardized food images and repeated ratings of two key constructs: liking (subjective preference) and perceived diabetes-related healthiness (belief about suitability for glycemic management). The tool was designed to be flexible and deployable across settings (e.g., clinics, laboratories, remote assessments), requiring minimal equipment and a short completion time.

We evaluated whether the tool performed as expected across several validation criteria. Specifically, we examined whether it was sensitive to changes by detecting intervention-related shifts in perceived healthiness and liking over time; whether it demonstrated internal consistency, such that theoretically related constructs (liking and perceived healthiness) changed in a coordinated manner; and whether it showed external validity, reflected by meaningful associations between tool-derived indices and external benchmarks, including expert dietitian ratings and physiological measures (e.g., indices of glucose stability). Finally, we assessed whether the tool differentiated between intervention contexts (e.g., monitoring modality) and captured item-level patterns related to food consumed by patients.

The tool was implemented within a broader clinical study examining whether short-term use of continuous glucose monitoring (CGM) improved outcomes in individuals with uncontrolled Type 2 diabetes mellitus (T2DM). Patients were assigned either to an intervention group using CGM for continuous glucose monitoring, or to a control group practicing self-monitoring of blood glucose (SMBG) via finger-prick testing. Patients in the CGM group had real-time access to glucose data via a smartphone application. For both the CGM and SMBG groups, glucose profiles were reviewed and discussed during follow-up visits with the medical team.

To the best of our knowledge, no prior research has directly examined the relationship between the glycemic response to specific foods and their perceived healthiness and desirability. Our framework further allowed tracking within-person changes in food evaluation as patients became more aware of health implications of their food choice, providing a framework to examine perceptual shifts alongside clinical or educational components of behavior change. While the broader clinical implications of CGM fall outside the scope of this study, our aim was to introduce a reliable and flexible method for capturing how health-focused interventions shape subjective food evaluations, an approach that may be applicable across nutrition, health psychology, and behavior change research.

Finally, we developed a prototype smartphone application to exemplify the experimental tool on a mobile interface (see https://nutri-shift-d8057cb0.base44.app). This application is provided for demonstration purposes only, to familiarize readers with the task’s structure and user experience; it was not used to generate the results reported here. We include it to illustrate the feasibility of self-administered, low-burden assessments that could be deployed at scale in subsequent research.

## Methods

### Patients

Fourteen patients (9 men, 5 women; mean age = 64.7 5 ± 9.45 years) were included in the study, based on the inclusion criteria of the parent clinical trial. Seven patients were assigned to the CGM group and seven to an SMBG control group. The study was approved by the Clinical Trials Department of the Israeli Ministry of Health (Helsinki approval no. SOR-0152-22). Participants’ privacy rights were kept throughout the study.

### Inclusion criteria

Eligible patients were adults (≥18 years) with uncontrolled T2DM (HbA1c ≥8.0%), who were willing and able to provide informed consent. Patients were required to receive therapy from all three major treatment classes: metformin, sodium-glucose cotransporter-2 inhibitors (SGLT-2i) and glucagon-like peptide-1 (GLP-1) receptor agonists, for at least 3 months prior to enrollment. Patients could also be enrolled if there was a contraindication to one or more of these medications, intolerance due to adverse effects, or ineligibility for treatment coverage under the national health services. Patients were required to be willing and able to use the study’s basic technology (e.g., smartphone applications and glucose-monitoring devices). Patients with or without basal insulin therapy were eligible.

### Exclusion criteria

Patients treated with multiple daily insulin injections (MDI) or a continuous subcutaneous insulin infusion, use of a CGM in the past 6 months, ongoing glucocorticoid therapy, pregnancy, and a life expectancy of less than 1 year.

### Design and stimuli

The study design and assessments timeline are summarized in Figure 1.

**Figure 1:**
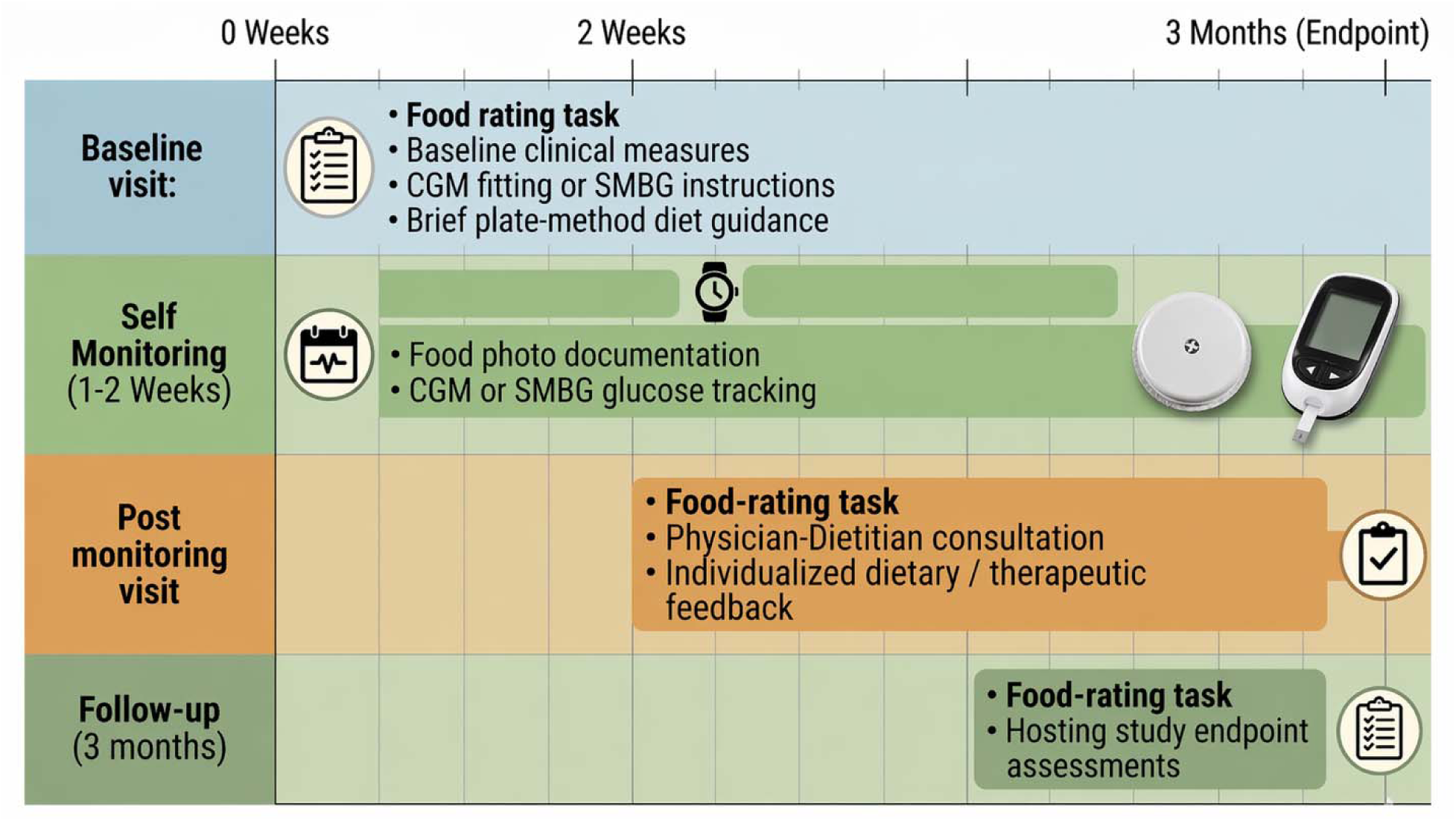
Study design. Patients attended three clinic visits, with a self-monitoring period conducted between the first and second visit. CGM: Continuous glucose monitoring, SMBG: self-monitoring of blood glucose.

#### First visit

At baseline, patients in both groups completed a computer-based food-rating task. Baseline clinical and laboratory characteristics were collected, including blood and urine tests, anthropometric measurements, and blood pressure. Patients in the intervention group were fitted with a CGM device, while control patients were instructed to perform SMBG via finger-prick testing four times daily (fasting and 2 postprandially). All patients received standardized dietary guidance emphasizing vegetables, protein, and healthy fats, with limited intake of processed foods and sugars.

#### Self-monitoring period (1-2 weeks)

During the self-monitoring period, patients tracked their glycemic responses to different foods by CGM (intervention group) or SMBG (control group). Patients documented food intake by photographing meals which were reviewed with the supervising physician to facilitate structured interpretation of patient dietary patterns in relation of glucose responses.

#### Post-monitoring assessment

Following the monitoring period, patients attended a consultation with a physician and a clinical dietitian, during which individualized therapeutic and dietary recommendations were provided based on recorded glucose responses. After this consultation, patients repeated the computer-based food-rating task.

#### Follow-up visit (3 months post-baseline)

At the follow-up visit, patients again completed the food-rating task for the third time. Clinical, laboratory, anthropometrics and SMBG data were collected as part of the hosting study’s predefined endpoint assessment.

### Food-rating task

To capture a broad range of food categories, 42 standardized food images (Shutterstock image base) were selected in collaboration with a clinical dietitian. The task comprised two rating sections, each including the same food items presented in a randomized order. In the preference-rating section, patients rated their subjective liking for each food item. In the health-perception section, patients rated how healthy each item was perceived to be in relation to their diabetes management (see Figure 2). A prototype mobile smartphone application (https://nutri-shift-d8057cb0.base44.app) was developed to demonstrate the functionality of the tool (this demonstration was not used for data collection).

**Figure 2:**
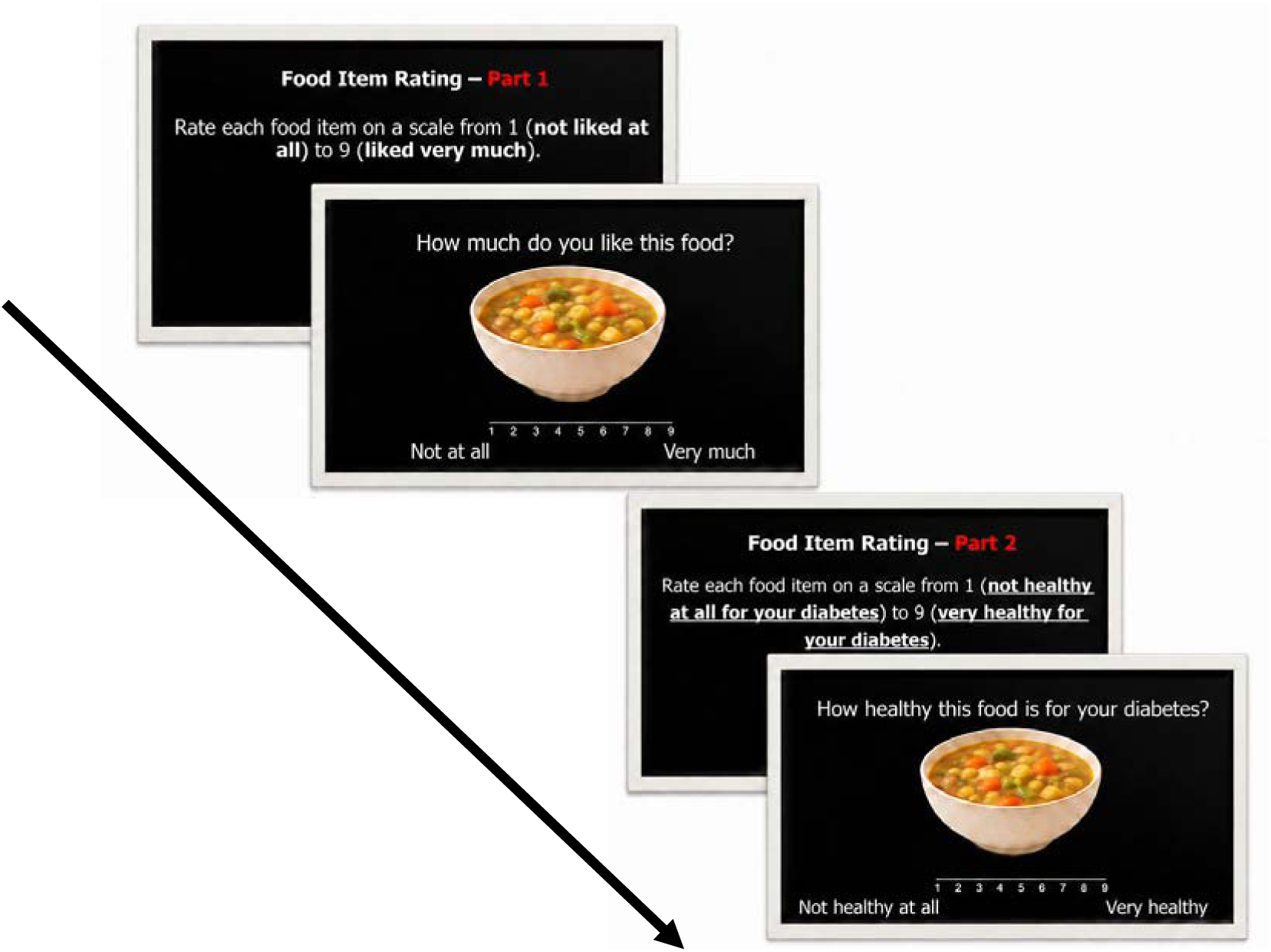
Illustration of the task, showing an example food-rating item.

Ratings of both preference and perceived healthiness were made on a scale of 1-9. Each food item was presented on the screen for 4 seconds, and the full task lasted approximately 8 minutes. At the beginning of each session, the experimenter recorded basic patient information, including study ID, age, and sex. Patients then reviewed standardized on-screen instructions and completed the task independently.

### Data analysis

All statistical analyses were conducted using Python (version 3.10.9), with scipy (Virtanen et al., 2020) package for statistical analysis. Spearman and Pearson correlations were used for correlation tests. Given the exploratory nature of our tool, we conducted several analyses to address key questions that emerged from the data (see Table 1 for summarized statistical tests and analysis).

**Table 1.** Analysis overview describing research questions and methodological tools used in the pilot.

| **Validation aim** | **Research question** | **Outcome measure(s)** | **Statistical approach** | **Interpretation** |
| --- | --- | --- | --- | --- |
| **Internal coherence** | Are liking and perceived healthiness correlated? | Mean liking and perceived healthiness ratings per session | Spearman correlation (levels and change scores) | Beliefs about health and taste rated in a coordinated manner |
| **Sensitivity to change** | Does the tool detect change over time? | Change scores in liking and perceived healthiness (V2-V1, V3-V1) | Spearman correlation of change scores | Tool captures intervention-related perceptual change |
| **External validity**<br>(*expert agreement*) | Do patients' ratings align with dietitian judgments? | R-score: correlation between patient and dietitian healthiness ratings (per visit); $\Delta R$-scores across visits | One-sample t-tests on $\Delta R$-scores | Tool captures change toward expert nutritional judgments |
| **External validity**<br>(**physiological relevance**) (CGM subgroup) | Is perceptual alignment associated with glucose stability? | R-score: correlation between changes in healthiness and liking (V2-V1); CGM coefficient of variation (CV) | Spearman correlation (R-score × CV) | Stronger perceptual coupling is associated with greater glucose stability |
| **Context sensitivity** | Does monitoring modality influence rating changes? | Change scores in liking and perceived healthiness | Mann-Whitney U tests (CGM vs. SMBG) | Monitoring context affects perceptual change |
| **Item-level sensitivity** | Are changes stronger for foods consumed? | Absolute change in ratings for consumed vs. all items | Wilcoxon signed-rank test | Tool is sensitive to personally relevant food exposure |

Abbreviations: V1-V3, visit 1-3; CV, coefficient of variation; CGM, continuous glucose monitoring; SMBG, self-monitoring of blood glucose; $\Delta R$ , delta R scores.

### Tool-derived indices and validation outcomes

To evaluate the tool as a methodological instrument, we derived six classes of outcomes aligned with the predefined validation aims:

#### 1. Internal coherence: coupling between liking and perceived healthiness

To assess internal coherence, we examined the association between liking and perceived healthiness ratings. Mean ratings for each construct were calculated across food items for each patient, and Spearman correlation analyses were conducted to evaluate the relationship between liking and perceived healthiness.

#### 2. Sensitivity to change over time

To examine whether the tool captures changes in food evaluation across the intervention, we computed change scores for both liking and perceived healthiness between baseline and the subsequent assessments (Visit 2 − Visit 1, Visit 3 − Visit 1). For each participant, mean change score was calculated across items, Spearman correlation analysis was then performed to assess whether changes in perceived healthiness were associated with concurrent changes in liking.

#### 3. External validity: agreement with dietitian ratings

Agreement between patients perceived healthiness rating and those provided by a clinical dietitian was quantified using an R-score, defined as the Pearson correlation between each patient’s ratings and the dietitian’s ratings across all food items at a given visit. R-scores were computed separately for each visit. Changes in agreement over time were calculated as Δ2 = R-score (Visit 2) − R-score (Visit 1) and Δ3 = R-score (Visit 3) − R-score (Visit 2). patient. One-sample t-tests were used to test whether these changes differed significantly from zero, indicating increased alignment with expert rating over time.

#### 4. External validity: association with physiological stability (CGM subgroup)

Within the CGM group, we examined whether alignment between perceptual changes was associated with patient glucose levels. For each patient, we calculated an R-score reflecting the correlation between changes in perceived healthiness and liking from visit 1 to visit 2 (i.e., following the glucose-monitoring period). Higher R-scores indicate stronger alignment between these perceptual changes. Glucose variability was quantified using the coefficient of variation (CV), calculated as the standard deviation divided by the mean glucose level derived from CGM data. Higher CV values indicated greater glucose variability (lower stability). Spearman correlation analysis was conducted to assess the relationship between CV and the R-score.

#### 5. Differentiation between intervention contexts

To examine whether exposure to CGM monitoring differentially influenced rating changes relative to the control conditions, changes in liking and perceived healthiness ratings were compared between the CGM and SMBG groups using Mann-Whitney U tests.

#### 6. Sensitivity to foods that the patient consumed during the intervention

To assess whether the tool was sensitive to foods that patients actually consumed during the monitoring period, we conducted a pilot item-level analysis based on food photographs documented by the patients. Consumed items were either directly matched to items in the task or mapped to the closest available equivalent (e.g. “bread with cheese spread” matched to “cheese bagel”). Absolute changes in liking and perceived healthiness ratings for consumed items were compared with changes across the full set of items using Wilcoxon signed-rank tests.

## Results

Table 3 summarizes the results according to the analysis plan, presenting the main estimates and statistical tests for each research question, along with brief interpretations.

### 1. Internal coherence: coupling between liking and healthiness

Mean ratings of perceived healthiness and liking were calculated across patients at each visit (Table 2). Spearman correlation analyses revealed a strong positive association between healthiness and liking at baseline, which remained significant across subsequent visits (Table 4). These findings indicate that foods perceived as healthier were also consistently rated as more liked, and that this relationship was stable over time.

**Table 2.**
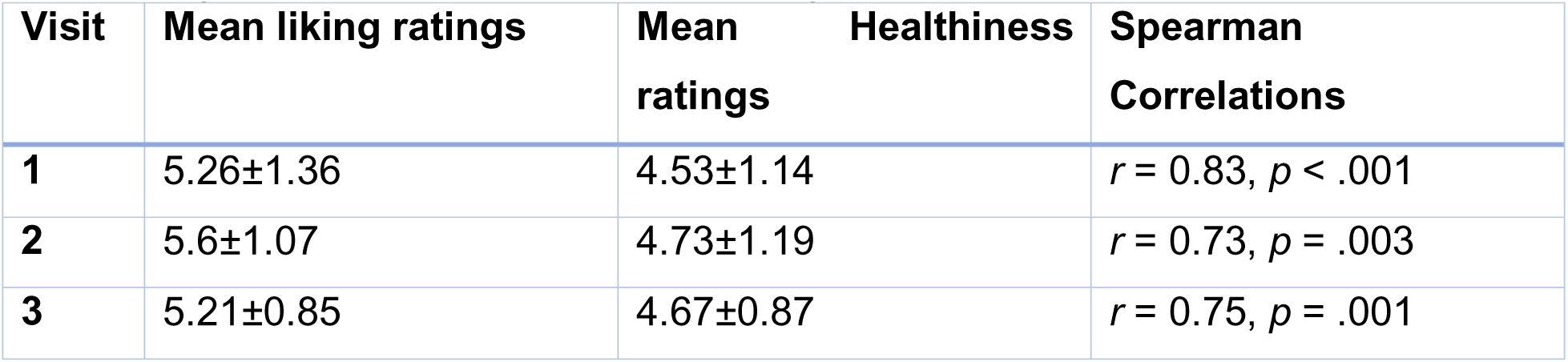
Mean ratings of perceived Healthiness and Liking across patients and their correlations.

| Visit | Mean liking ratings | Mean Healthiness ratings | Spearman Correlations |
| --- | --- | --- | --- |
| 1 | 5.26±1.36 | 4.53±1.14 | $r = 0.83, p < .001$ |
| 2 | 5.6±1.07 | 4.73±1.19 | $r = 0.73, p = .003$ |
| 3 | 5.21±0.85 | 4.67±0.87 | $r = 0.75, p = .001$ |

**Table 3.** A summary of the results according to the analysis plan.

| **Research question** | **Outcome measure** | **Statistical test** | **Key results** | **Interpretation** |
| --- | --- | --- | --- | --- |
| **1. Internal coherence: coupling between liking and healthiness** | Mean liking and healthiness ratings per visit | Spearman correlation | Strong positive correlations at all visits (Visit 1: $r = 0.83$, $p < .001$; Visit 2: $r = 0.73$, $p = .003$; Visit 3: $r = 0.75$, $p = .001$) | Healthiness and liking ratings were consistently aligned across sessions |
| **2. Sensitivity to change over time** | Change scores (Visit 2-1; Visit 3-1) | Spearman correlation | Significant coupling of changes at Visit 2 ($r = 0.76$, $p = .001$) and Visit 3 ($r = 0.64$, $p = .012$) | Changes in liking and healthiness tended to occur in tandem |
| **3. External validity: agreement with dietitian ratings** | R-score (patient-dietitian correlation per visit) and change in R-scores ($\Delta 2$, $\Delta 3$) | One-sample t-tests | Trend toward increased alignment from Visit 1 to Visit 2 ($\Delta 2$: $t(13) = 1.89$, $p = .081$); no change thereafter ($\Delta 3$: $p = .174$) | Ratings showed increased alignment with expert judgments after monitoring |
| **4. External validity: association with physiological stability (CGM subgroup)** | Correlation between R-score (change coupling) and glucose CV | Spearman correlation | Significant negative association ($r = -0.821$, $p = .023$, $N = 7$) | Greater glucose stability was associated with stronger health-liking coupling |
| **5. Discriminant sensitivity across intervention contexts (CGM vs. SMBG)** | Change scores in ratings (Visit 2-1; Visit 3-1) | Mann-Whitney U test | No significant group differences (all $p > .05$) | No evidence that monitoring modality altered rating changes in this pilot |
| **6. Item level Sensitivity** | Absolute change in ratings for consumed vs. all items | Wilcoxon signed-rank test | Smaller changes for consumed items ($\Delta = 0.73 \pm 0.57$) compared to all items ($\Delta = 1.27 \pm 0.62$; $W = 20$, $p = .04$) | Consumed items showed smaller rating shifts |

Abbreviations: CV, coefficient of variation; CGM, continuous glucose monitoring; SMBG, self-monitoring of blood glucose; $\Delta R$ , delta R scores.

**Table 4.** Descriptive statistics of alignment between patients’ and the dietitian’s healthiness ratings.

|  | Mean difference in food healthiness ratings | Mean R-scores |
| --- | --- | --- |
| Visit 1 | $0.08 \pm 1.98$ | $0.199 \pm 0.18$ |
| Visit 2 | $0.27 \pm 2.04$ | $0.29 \pm 0.13$ |
| Visit 3 | $0.22 \pm 2.24$ | $0.25 \pm 0.17$ |

### 2. Sensitivity to change

From the first to the second visit, mean ratings increased by 0.34 ± 1.01 for liking and by 0.19 ± 0.80 for perceived healthiness, with substantial inter-patient variability. When comparing the third visit with the first, liking ratings showed a slight decrease (*M* = -0.04 ± 1.09), whereas healthiness ratings showed a modest increase (*M* = 0.13 ± 0.98), again with considerable variability across patients. Overall, liking ratings exhibited greater temporal variability, whereas healthiness ratings showed a more consistent pattern across sessions. Mean differences between visits were not statistically significant.

To assess whether the tool captured coordinated change between subjective preference and health perception, we examined correlation between changes in liking and healthiness ratings. Spearman correlation analysis revealed a strong positive association between changes from the first to second visit (*r* = 0.76, *p* = .001; Figure 3A), as well as from the first to the third visit (*r* = 0.64, *p* = .012; Figure 3B). These findings indicate that patients who showed increased (or decreased) in liking tended to exhibit corresponding changes in perceived healthiness. No significant differences were observed in the magnitude of change across visits.

**Figure 3.**
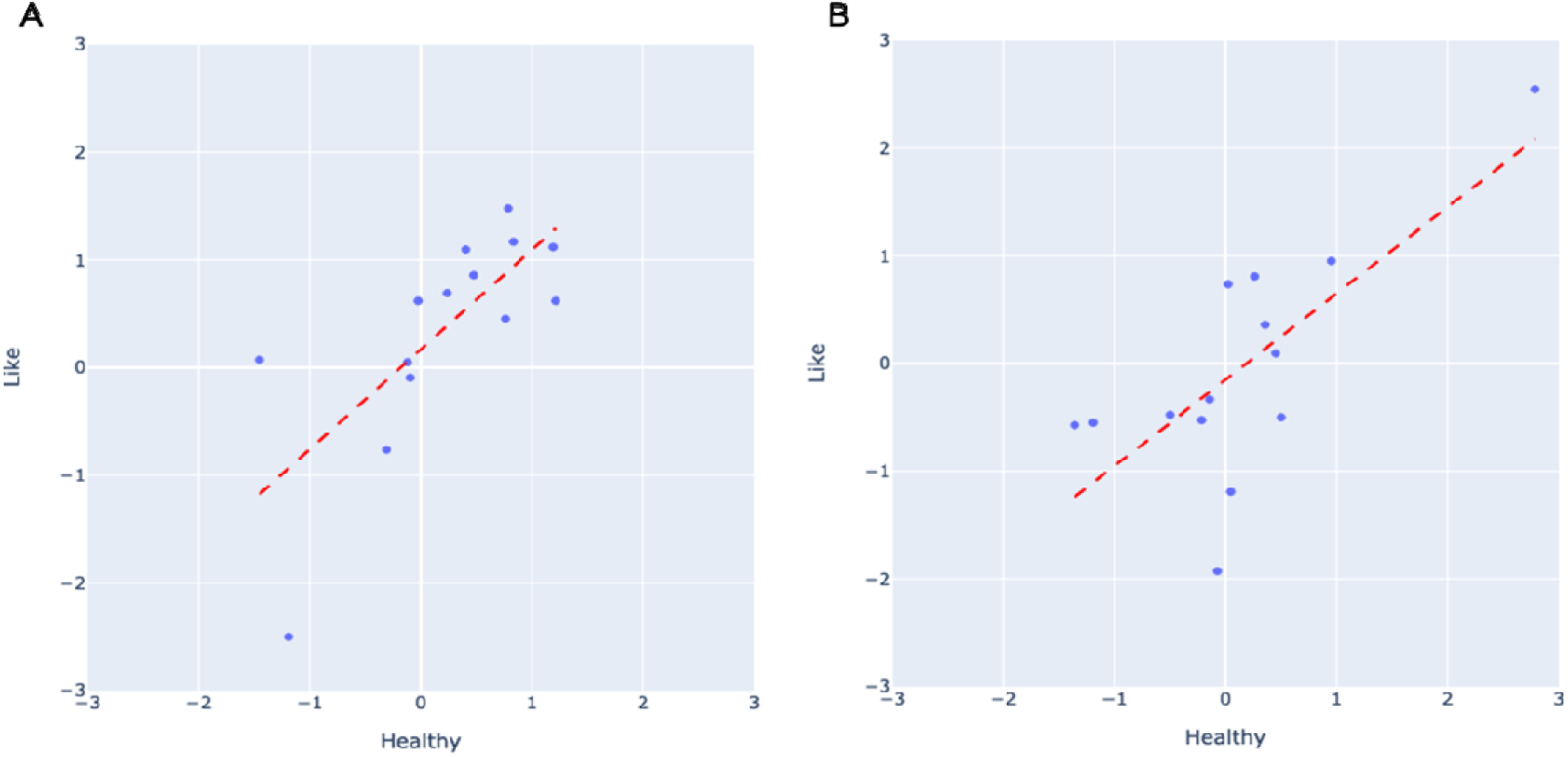
Mean score of differences in ratings between the first visit and subsequent visits. A. Second visit, *r* = 0.76, *p* = .001. B. Third visit, *r* = 0.64, *p* = .012.

### 3. External validity: agreement with dietitian ratings

The mean difference in food item healthiness ratings between patients and the dietitian at the first visit (Table 4) indicated that, on average, patients and the dietitian rated items similarly, albeit with substantial inter-patient variability. At the second and third visits, mean differences increased slightly and remained highly variable across patients. However, differences between visits were not statistically significant.

In contrast, analysis based on R-scores, quantifying the correspondence between patients’ healthiness rating and the dietitian’s rating across items, revealed a trend towards increased alignment following the monitoring period (Δ2: *t*(13) = 1.89, *p* = .081, *M* = 0.09±0.18). The changes in R-scores between the second and third visits (Δ3) was not significant (*t*(13) = −1.44, *p* = .174, *M* = -0.04±0.11), suggesting that alignment remained relatively stable thereafter (Table 4).

Overall, despite patients providing higher absolute healthiness ratings than the dietitian, the relative pattern of ratings across food items became more closely aligned with the dietitian’s judgment following the intervention.

### 4. External validity: association with physiological stability (CGM subgroup)

Because the coefficient of variation (CV) reflects glucose variability normalized to each patient’s mean glucose level, it enables comparisons of glycemic stability across patients despite differences in baseline glycemia and is considered a robust index of glucose variability. In the CGM subgroup, Correlation analysis examining the association between glucose stability (CV) and R-score, reflecting the coupling between first-to-second visit changes in liking and healthiness ratings, revealed a strong negative relationship (*r* = -0.82, *p* = .023). This preliminary result indicates that patients with more stable glucose profiles (lower CV) demonstrated stronger concordance between changes in liking and perceived healthiness ratings (Figure 4).

**Figure 4.**
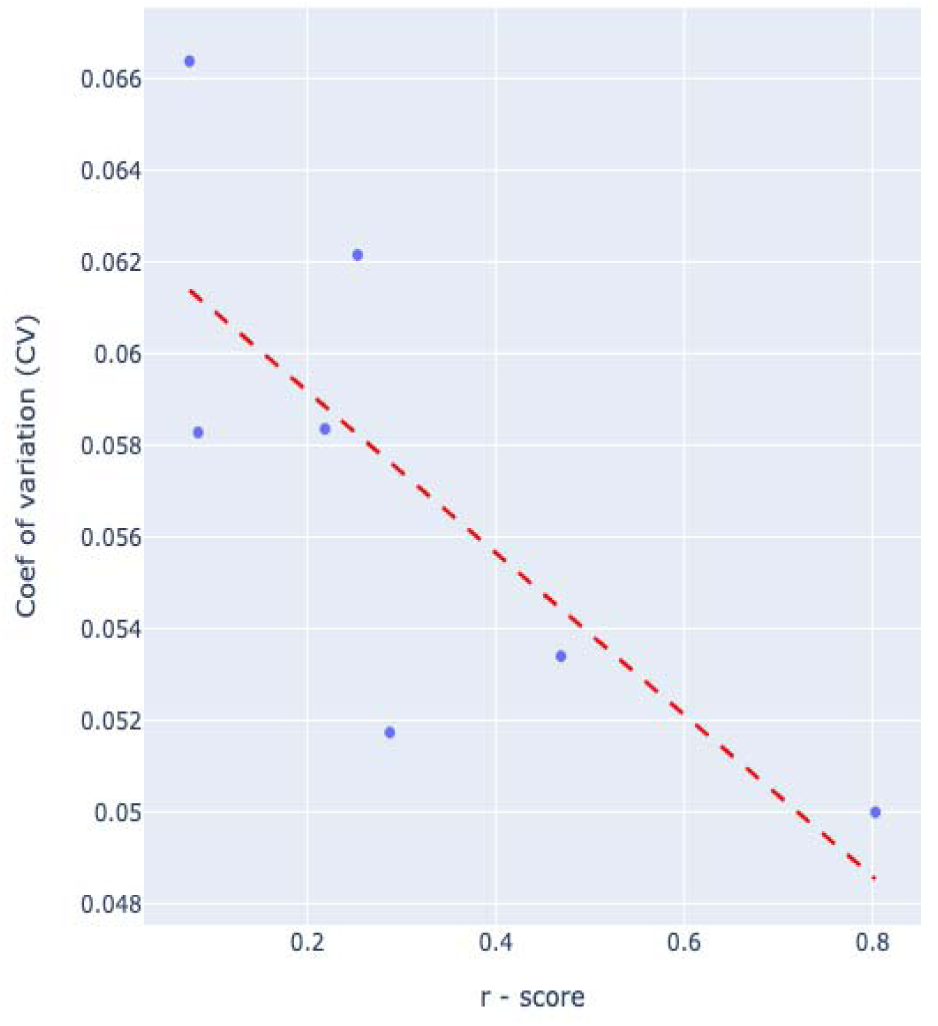
Correlation results for r-score, calculated from the correlation of the change in healthiness and liking ratings from the first visit to the second visit, and the coefficient of variation (CV) of each patient. A significant negative relationship between CV and the r-score was found (*r* = -0.821, *p* = .023).

### 5. Discriminant sensitivity across intervention contexts

To examine whether the tool differentiates between intervention contexts with differing levels of glucose monitoring, we compared changes in liking and perceived healthiness ratings between the CGM and SMBG groups. No statistically significant group differences were observed at either the second or third visit for either rating dimension (Table 5).

**Table 5.** Between-group comparisons of changes in food liking and perceived healthiness ratings (CGM vs. SMBG)

| Visit | Test | CGM group (n=7) | SMBG group (n=7) | Mann-Whitney result |
| --- | --- | --- | --- | --- |
| **2<sup>nd</sup> Visit** | “Like” Mean difference | 0.69±0.72 | 0.00±1.18 | $U = 14.5$,<br>$p = .22$ |
| | “Healthiness” Mean difference | 0.46± 0.5 | -0.08±0.97 | $U = 17$,<br>$p = .38$ |
| **3<sup>rd</sup> Visit** | “Like” Mean difference | 0.32±1.16 | -0.41±0.93 | $U = 17$,<br>$p = .38$ |
| | “Healthiness” Mean difference | 0.36±1.28 | -0.09±0.57 | $U = 21$,<br>$p = .71$ |

### 6. Item-level sensitivity: Does the tool detect differential rating changes for foods directly experienced during the intervention?

The number of food items documented by patients during the monitoring period were relatively small (mean 4.21±1.84 items). To examine item-level sensitivity, we computed changes in perceived healthiness ratings for foods that patients consumed during the intervention with changes observed across the full item set. Results indicated that the mean change in healthiness rating at the second visit was smaller for consumed items (*M* = 0.73 ± 0.57) than for all items combined (*M* = 1.27 ± 0.62). This difference reached statistical significance (*W* = 20, *p* = .04), suggesting that the tool is sensitive to differential rating patterns at the item level.

## Discussion

In this pilot study, we developed and tested a research tool designed to assess changes in food preferences and perceived diabetes-related healthiness within the context of a medically supervised nutritional intervention. An online demonstration version of the tool is available at https://nutri-shift-d8057cb0.base44.app. The tool was embedded in a larger study comparing real-time continuous glucose monitoring (CGM) and self-monitoring of blood glucose (SMBG) among individuals with T2DM. Consistent with the methodological aim of this manuscript, the primary objective was not to evaluate clinical efficacy, but to examine the tool’s performance across six validation criteria. Overall, the findings suggest that the tool yields coherent, interpretable indices of food-related perception that are sensitive to within-person dynamics and show preliminary links to external benchmarks.

### Internal coherence and sensitivity to change

As a first validation criterion, we examined internal coherence between the two rating dimensions. The tool demonstrated strong internal coherence, with robust positive correlations between liking and perceived healthiness across all visits. The stability of this association across repeated administrations indicates that the tool consistently captures a structured relationship between evaluative dimensions, supporting its reliability as a measure of food-related perceptions over time.

### External validity: expert alignment and physiological relevance

A key validation criterion was whether perceived healthiness rating converged toward expert judgment and physiological indicators. We observed a trend-level increase in alignment between patients’ and dietitian’s healthiness ratings from Visit 1 to Visit 2, followed by relative stability thereafter. This pattern suggests that the tool can detect gradual convergence in the relative structure of ratings, even when absolute scale use differs between raters. Such convergence supports the tool’s external validity as a measure of health-related food perception.

Although no differences in rating changes were observed between CGM and SMBG groups, within the CGM subgroup lower glucose variability (indexed by coefficient of variation) was significantly associated with coupling between changes in liking and healthiness ratings. This association provides preliminary external validation, indicating that tool-derived indices related meaningfully to independent physiological measures. Importantly, this finding does not imply causality but suggests that the tool captures perceptual dynamics that co-vary with physiological stability.

### Item-level sensitivity (consumed vs non-consumed foods)

The tool also demonstrated item-level sensitivity. Changes in perceived healthiness were smaller for foods documented as consumed during the intervention compared with the full stimulus set. This finding suggests that the tool can distinguish between foods directly experienced during the intervention and those evaluated more abstractly. Methodologically, this result indicates that the tool is sensitive to differential exposure while also highlighting limits in belief updating over short intervention periods.

### Limitations and future directions

Several limitations should be considered when interpreting these findings. First, the absence of a non-intervention control condition limits the ability to disentangle tool sensitivity from general study participation effects. Second, the small sample size restricts statistical power and limits generalizability. In addition, the short monitoring period may not be sufficient to induce large or stable perceptual shifts, emphasizing the need for longer-term validation studies.

Finally, reliance on self-documented food intake constrained analyses linking ratings to actual consumption, underscoring the importance of integrating more objective dietary tracking methods in future work.

### Conclusions

This pilot study introduces a brief, low-burden tool for repeatedly assessing food liking and perceived diabetes-related healthiness in the context of nutrition-focused interventions. In a pilot implementation, the tool demonstrated internal coherence, sensitivity to within-person change patterns and preliminary correspondence with expert and physiological benchmarks. These properties suggest that the tool may support intervention research by enabling scalable quantification of perceptual change, an important intermediate construct through which interventions may influence dietary behavior. Future studies should validate the tool in larger samples, formally assess its measurement properties, and examine its utility for predicting longer-term behavioral and health outcomes.

## Supporting information

Image list

## Data Availability

All data produced are available online at

https://github.com/SchonbergLab/Food-preferences-diabetes.git

## Declaration of generative AI and AI-assisted technologies in the writing process

During the preparation of this work the authors used ChatGPT to enhance content and refine text. After using this tool, the authors reviewed and edited the content as needed and take full responsibility for the content of the published article.

The demo application was developed using Base44 and is provided solely for demonstration purposes, without representing a finalized research tool.

## Data availability

Data is available at the Schonberg-lab GitHub at: https://github.com/SchonbergLab/Food-preferences-diabetes.

## References

Bédard, A., Lamarche, P.-O., Grégoire, L.-M., Trudel-Guy, C., Provencher, V., Desroches, S., & Lemieux, S. (2020). Can eating pleasure be a lever for healthy eating? A systematic scoping review of eating pleasure and its links with dietary behaviors and health. Plos One, 15(12), e0244292. 10.1371/journal.pone.0244292

Berry, C., & Romero, M. (2021). The fair trade food labeling health halo: Effects of fair trade labeling on consumption and perceived healthfulness. Food Quality and Preference, 94, 104321. 10.1016/j.foodqual.2021.104321

Burch, E., Ball, L., Somerville, M., & Williams, L. T. (2018). Dietary intake by food group of individuals with type 2 diabetes mellitus: A systematic review. Diabetes Research and Clinical Practice, 137, 160–172. 10.1016/j.diabres.2017.12.016

Dalton, M., Blundell, J., & Finlayson, G. S. (2013). Examination of food reward and energy intake under laboratory and free-living conditions in a trait binge eating subtype of obesity. Frontiers in Psychology, 4, 757. 10.3389/fpsyg.2013.00757

Dobrow, L., Estrada, I., Burkholder-Cooley, N., & Miklavcic, J. (2021). Potential effectiveness of registered dietitian nutritionists in healthy behavior interventions for managing type 2 diabetes in older adults: A systematic review. Frontiers in Nutrition, 8, 737410. 10.3389/fnut.2021.737410

Durand, F., Besson, T., Flaudias, V., & Zerhouni, O. (2025). New challenges and perspectives on the organic halo effect on calories estimation: A brief review and Meta-analysis. Food Quality and Preference, 127, 105460. 10.1016/j.foodqual.2025.105460

Ewers, B., Sørensen, M. R., Fagt, S., Diaz, L. J., & Vilsbøll, T. (2021). Intention and Perceptions of Healthy Eating versus Actual Intake Among Patients with Type 1 and Type 2 Diabetes and the General Population. Patient Preference and Adherence, 15, 2027–2037. 10.2147/PPA.S325214

Falk, A., & Zimmermann, F. (2017). Information processing and commitment. The Economic Journal, 128(613), 1983–2002. 10.1111/ecoj.12542

Gal, A. M., Iatcu, C. O., Popa, A. D., Arhire, L. I., Mihalache, L., Gherasim, A., Nita, O., Soimaru, R. M., Gheorghita, R., Graur, M., & Covasa, M. (2024). Understanding the interplay of dietary intake and eating behavior in type 2 diabetes. Nutrients, 16(6). 10.3390/nu16060771

Gardner, B., & Lally, P. (2018). Modelling habit formation and its determinants. In B. Verplanken (Ed.), The psychology of habit: theory, mechanisms, change, and contexts (pp. 207-229). Springer International Publishing. 10.1007/978-3-319-97529-0_12

Guideline, N. (2022). Type 2 diabetes in adults: Management (update). National Institute for Health and Care Excellence. https://www.ncbi.nlm.nih.gov/books/NBK580244/bin/niceng28er3_bm1.pdf

Haasova, S., & Florack, A. (2019). Practicing the (un)healthy = tasty intuition: Toward an ecological view of the relationship between health and taste in consumer judgments. Food Quality and Preference, 75, 39–53. 10.1016/j.foodqual.2019.01.024

Han, C. Y., Chan, C. G. B., Lim, S. L., Zheng, X., Woon, Z. W., Chan, Y. T., Bhaskaran, K., Tan, K. F., Mangaikarasu, K., & Chong, M. F.-F. (2020). Diabetes-related nutrition knowledge and dietary adherence in patients with Type 2 diabetes mellitus: A mixed-methods exploratory study. Proceedings of Singapore Healthcare, 29(2), 81–90. 10.1177/2010105820901742

Lange, R. D., Chattoraj, A., Beck, J. M., Yates, J. L., & Haefner, R. M. (2021). A confirmation bias in perceptual decision-making due to hierarchical approximate inference. PLoS Computational Biology, 17(11), e1009517. 10.1371/journal.pcbi.1009517

Martens, T. W., Willis, H. J., Bergenstal, R. M., Kruger, D. F., Karslioglu-French, E., & Steenkamp, D. W. (2025). A Randomized Controlled Trial Using Continuous Glucose Monitoring to Guide Food Choices and Diabetes Self-Care in People with Type 2 Diabetes not Taking Insulin. Diabetes Technology & Therapeutics, 27(4), 261–270. 10.1089/dia.2024.0579

Nadricka, K., Millet, K., & Verlegh, P. W. J. (2020). When organic products are tasty: Taste inferences from an Organic = Healthy Association. Food Quality and Preference, 83, 103896. 10.1016/j.foodqual.2020.103896

Peng, M., Browne, H., Cahayadi, J., & Cakmak, Y. (2021). Predicting food choices based on eye-tracking data: Comparisons between real-life and virtual tasks. Appetite, 166, 105477. 10.1016/j.appet.2021.105477

Pot, G. K., Battjes-Fries, M. C., Patijn, O. N., Pijl, H., Witkamp, R. F., de Visser, M., van der Zijl, N., de Vries, M., & Voshol, P. J. (2019). Nutrition and lifestyle intervention in type 2 diabetes: pilot study in the Netherlands showing improved glucose control and reduction in glucose lowering medication. *BMJ Nutrition*, Prevention & Health, 2(1), 43–50. 10.1136/bmjnph-2018-000012

Previdelli, Á. N., de Andrade, S. C., Fisberg, R. M., & Marchioni, D. M. (2016). Using Two Different Approaches to Assess Dietary Patterns: Hypothesis-Driven and Data-Driven Analysis. Nutrients, 8(10). 10.3390/nu8100593

Raghunathan, R., Naylor, R. W., & Hoyer, W. D. (2006). The unhealthy = tasty intuition and its effects on taste inferences, enjoyment, and choice of food products. Journal of Marketing, 70(4), 170–184. 10.1509/jmkg.70.4.170

Richetin, J., Demartini, E., Gaviglio, A., Ricci, E. C., Stranieri, S., Banterle, A., & Perugini, M. (2021). The biasing effect of evocative attributes at the implicit and explicit level: The tradition halo and the industrial horn in food products evaluations. Journal of Retailing and Consumer Services, 61, 101890. 10.1016/j.jretconser.2019.101890

Santos-Báez, L. S., Díaz-Rizzolo, D. A., Popp, C. J., Shaw, D., Fine, K. S., Altomare, A., St-Onge, M.-P., Manoogian, E. N. C., Panda, S., Cheng, B., & Laferrère, B. (2024). Diet and Meal Pattern Determinants of Glucose Levels and Variability in Adults with and without Prediabetes or Early-Onset Type 2 Diabetes: A Pilot Study. Nutrients, 16(9). 10.3390/nu16091295

Schwingshackl, L., Hoffmann, G., Lampousi, A.-M., Knüppel, S., Iqbal, K., Schwedhelm, C., Bechthold, A., Schlesinger, S., & Boeing, H. (2017). Food groups and risk of type 2 diabetes mellitus: a systematic review and meta-analysis of prospective studies. European Journal of Epidemiology, 32(5), 363–375. 10.1007/s10654-017-0246-y

Siopis, G., Colagiuri, S., & Allman-Farinelli, M. (2021). Effectiveness of dietetic intervention for people with type 2 diabetes: A meta-analysis. Clinical Nutrition, 40(5), 3114–3122. 10.1016/j.clnu.2020.12.009

Thorndike, E. L. (1920). A constant error in psychological ratings. Journal of Applied Psychology, 4(1), 25–29. 10.1037/h0071663

Virtanen, P., Gommers, R., Oliphant, T. E., Haberland, M., Reddy, T., Cournapeau, D., Burovski, E., Peterson, P., Weckesser, W., Bright, J., van der Walt, S. J., Brett, M., Wilson, J., Millman, K. J., Mayorov, N., Nelson, A. R. J., Jones, E., Kern, R., Larson, E., … SciPy 1.0 Contributors. (2020). SciPy 1.0: fundamental algorithms for scientific computing in Python. Nature Methods, 17(3), 261-272. 10.1038/s41592-019-0686-2

Vogel, E., & Mol, A. (2014). Enjoy your food: on losing weight and taking pleasure. In S. Cohn (Ed.), From health behaviours to health practices (pp. 145–157). Wiley. 10.1002/9781118898345.ch13

Werle, C. O. C., Trendel, O., & Ardito, G. (2013). Unhealthy food is not tastier for everybody: The “healthy=tasty” French intuition. Food Quality and Preference, 28(1), 116–121. 10.1016/j.foodqual.2012.07.007

