## Supplementary material for "A tool for assessing changes in food preferences and health perceptions during nutritional interventions": Image list

Food items rated in the task:

1. white_beans.png
2. Wafers.png
3. vegetables_soup.png
4. vegetables_salad2.png
5. vanilla_ice_cream.png
6. tuna_sandwich.png
7. tomato.png
8. Stuffed_Vine.png
9. stuffed_peppers.png
10. Spaghetti_with_tomato_sauce.png
11. shawarma.png
12. pizza.png
13. pepper.png
14. peas.png
15. pastrami.png
16. Omellete_bread.png
17. mixed_nuts.png
18. milk_glass.png
19. Lentil_Soup.png
20. labaneh.png
21. humus with pita.png
22. Hamburger.png
23. grilled_cheese_sandwich.png
24. grapes.png
25. french_fries.png
26. drumstick with potatos.png
27. daysa.png*
28. dates_cookies(ma_amoul).png
29. date fruit with walnut.png
30. Couscous.png**
31. corncob.png
32. chicken_breast_schnitzel.png
33. cheese_cake.png
34. cheese_bagel.png
35. cereals.png
36. burekas.png
37. bun.png
38. bread with avocado spread.png
39. beef stew on mashed potatos.png
40. banana.png
41. Apple.png
42. antipasti vegetables.png

*Daysa - Nillerdk - CC BY 3.0, <https://commons.wikimedia.org/w/index.php?curid=4070068>

**Couscous - Rainer Zenz, CC BY-SA 3.0 <http://creativecommons.org/licenses/by-sa/3.0/>, via Wikimedia Commons
